# Clinical Utility and Outcomes of Targeted Next-Generation Sequencing in Pediatric Respiratory Tract Infections: A Nine-Month Retrospective Study at a North Chinese Hospital

**DOI:** 10.1101/2024.09.18.24313214

**Authors:** Lixiang Wang, Hui Zhong, Lexia Yang, Xue Yang

**Author notes:** Correspondence: Lexia Yang,; Xue Yang,.

## Abstract

**Background/Objectives:** Children are at increased risk for respiratory tract infections (RTIs) due to their developing pulmonary functions and immune systems, highlighting the necessity of accurate and rapid pathogen diagnostic methods.

**Methods:** In this study, 940 pediatric hospitalized patients with RTIs were included to evaluate the clinical utility and outcomes of 107-pathogen targeted next-generation sequencing (tNGS) panel by comparing it with CMTs.

**Results:** Our study demonstrated that tNGS exhibited significantly higher sensitivity in pathogen detection compared to CMTs, with a detection rate of 91.38%. tNGS was capable of identifying a broader range of pathogens, including low-abundance and RNA viruses frequently missed by CMTs, and it was able to detect mixed infections, whereas CMTs typically identified only a single pathogen. Treatment adjustments based on tNGS results led to clinical improvements in the majority of patients, with 35.32% experiencing escalated treatment plans and 29.04% experiencing de-escalated treatment plans. Additionally, the analysis revealed significant variations in pathogen prevalence across different age groups and seasons, highlighting the need for tailored diagnostic, treatment, and prevention strategies.

**Conclusions:** The findings highlight the potential of tNGS to improve clinical outcomes in pediatric RTIs by providing more accurate and comprehensive pathogen detection.

## 1. Introduction

Respiratory tract infections (RTIs) in children are a critical and challenging issue due to their developing pulmonary function and relatively weaker immune resistance [1, 2]. Compared to adults, pediatric respiratory infections carry an increased risk of serious complications and long-term impacts. Statistics show that 7% to 13% of pediatric pneumonia cases can progress to severe pneumonia, imposing a heavy burden on families [3–5]. Furthermore, children have higher exposure to pathogens in communal environments like schools and daycare centers, making respiratory infections difficult to control and significantly increasing their prevalence and recurrence in this population [6]. Therefore, the rapid and accurate diagnosis of pathogens and epidemiologic tracing are crucial for effective pediatric patient care.

Conventional microbiological tests (CMTs), including microbial culture, antigen-antibody detection, and PCR assays, remain widely used and recognized methods for diagnosing pediatric respiratory infections but present challenges due to their limitations [7, 8]. Only a small fraction of pathogens can be cultured [9], antigen detection tests often have lower sensitivity [10], and PCR assays require specific prior knowledge to select the appropriate targets, resulting in difficulty detecting atypical pathogens and high false-negative rates. To overcome these limitations, metagenomic next-generation sequencing (mNGS) has emerged as a rapid and unbiased approach that allows for one test to identify all potential pathogens in a sample, making it a promising tool for clinical diagnosis. Previous studies have shown that mNGS has higher sensitivity in pathogen detection compared to CMTs and has demonstrated its advantages in pediatric respiratory infections [11–14], especially for uncommon pathogens. However, mNGS is also limited by variable sensitivity depending on specimen type and disease severity, high cost, and the challenge of distinguishing true pathogens from commensal microbes due to its comprehensive coverage of all microorganisms [15–17].

Probe capture-based targeted next-generation sequencing (tNGS) can enrich regions of interest pathogens and integrate the advantages of mNGS, improving the proportion of target reads, processing speed, and cost-efficiency. The tNGS approach has demonstrated its advantages in clinical pathogen discovery for blood, joint, and respiratory infections, with higher sensitivity in detecting specific low-abundance pathogens compared to mNGS and CMTs [18, 19]. For example, the application of tNGS in diagnosing pulmonary infections showed higher sensitivity and accuracy in identifying *Mycobacterium tuberculosis* and *nontuberculous mycobacteria*, making it a valuable tool in clinical settings for respiratory infections [20]. It is also able to detect drug-resistance genes, aiding in appropriate treatment decisions [21]. However, similar to mNGS method, the sensitivity of tNGS varies depending on the specimen and disease type, and the application of tNGS method in pediatric respiratory infections requires for further standardization of methodologies and interpretation criteria to ensure consistent and accurate results.

To date, tNGS pathogen detection applied for pediatric respiratory infections predominantly utilizes bronchoalveolar lavage fluid and is limited by the small scale of patient cohorts, with insufficient research on other sample types and large-scale populations [22–24]. In the present study, throat swab samples were collected from respiratory infected children from a hospital in northern China for nine months, and a 95-pathogen tNGS panel was applied in pathogen discovery to evaluate the impact of tNGS data on clinical outcomes by comparing with CMTs. Our analysis revealed the important role of the tNGS platform in respiratory pathogen identification in children, which may provide an optimal way to diagnose suspected pediatric respiratory infection disease.

## 2. Materials and methods

### 2.1 Patients enrolment and data collection

This retrospective study was conducted at Qingdao Huangdao District People’s Hospital, located in northern China. Children hospitalized from April 1, 2023, to December 31, 2023, in the pediatric department with respiratory tract infections (RTI) who underwent tNGS analysis were screened (**Figure 1**). Basic clinical data were collected from electronic medical records, including age, gender, and hospitalization duration. During the hospitalization period, data on complete blood counts, traditional pathogen detection, tNGS results, imaging findings, and treatment regimens were recorded.

**Figure 1.**
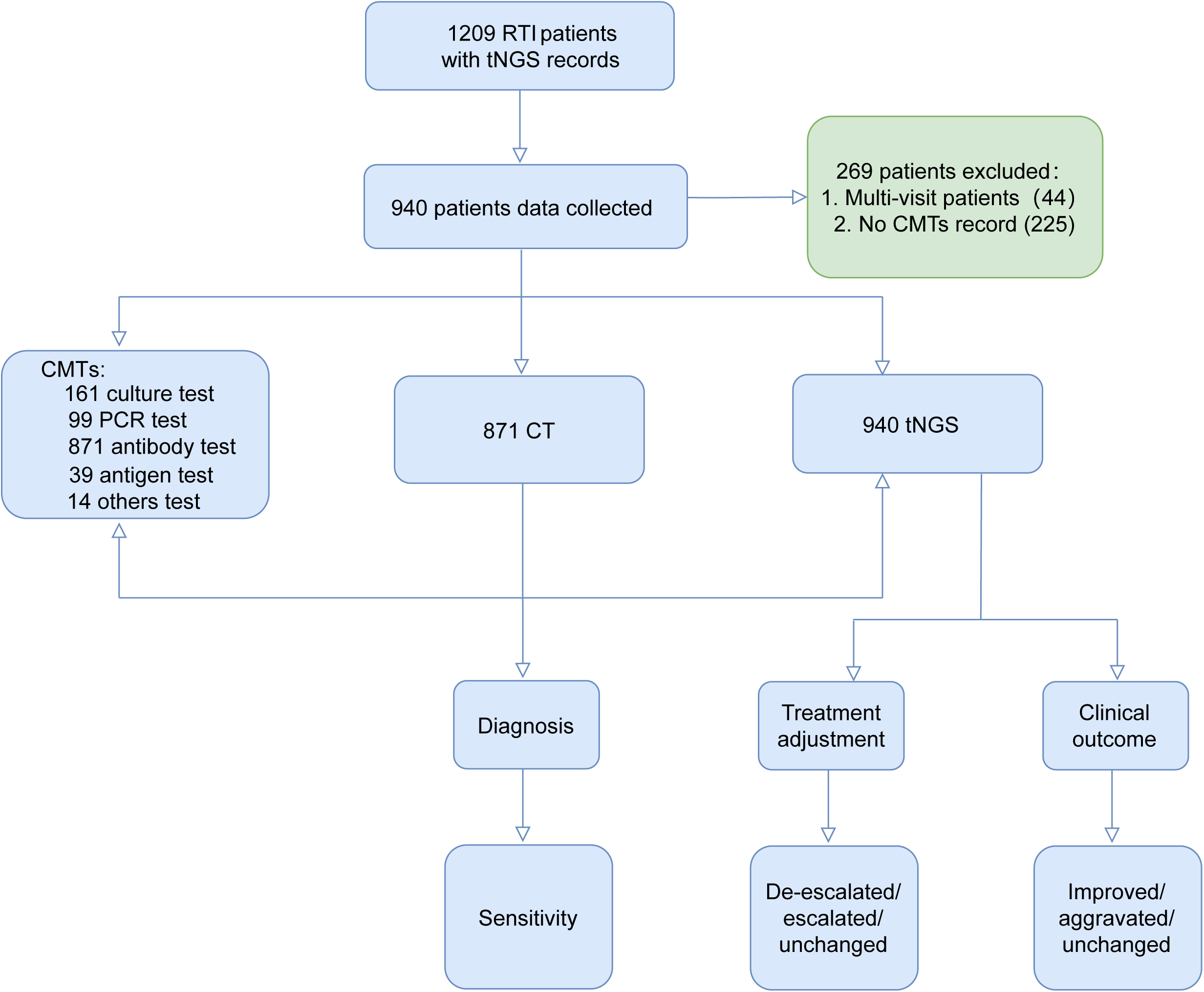
Flowchart of patient recruitment and sample collection. A total of 1,209 patients with suspected RTI and having tNGS test were screened from April 1, 2023, to December 31, 2023, at Qingdao Huangdao District People’s Hospital, Qingdao, China, and 940 were enrolled in the study. The clinical outcomes of tNGS were evaluated and compared with CMTs and CT. RTI, respiratory tract infection; CT, computed tomography; tNGS, targeted next-generation sequencing.

The diagnosis of RTI was based on a comprehensive assessment that combined clinical symptoms, epidemiological history, and pathogenetic or serological tests. Patients were considered for inclusion if they exhibited local symptoms such as nasal congestion, sneezing, runny nose, throat discomfort, sore throat, cough, sputum production, shortness of breath, wheezing, or hoarseness. Systemic symptoms including fever, fatigue, headache, restlessness, general malaise, lethargy, lack of appetite, vomiting, diarrhea, or abdominal pain were also considered. Common signs like pharyngeal congestion, tonsillar enlargement, cervical lymphadenopathy, tachypnea, cyanosis, and lung rales were taken into account. Furthermore, patients had to have at least one positive pathogenetic or serological test result that matched their clinical presentation, including a positive pathogen from the PCR, antigen, and culture tests, specific IgM in the convalescent phase, or a fourfold or greater increase in IgG antibody levels in the acute phase.

Patients were excluded from the study if they failed to provide informed consent, had multiple visits to the hospital, or had incomplete medical records. This study was conducted following the Declaration of Helsinki. The study protocol was approved by the Ethics Committee of Qingdao Huangdao District People’s Hospital (LL-LW-2024001). Written informed consent was obtained from all adult participants or their legal guardians for participation in this study.

### 2.2 Sample processing and conventional microbiological tests (CMTs)

Conventional microbiological tests (CMTs) in this study included microbial culture, antigen and antibody test, PCR test, *Cryptococcus neoformans* ink stain microscopy, and (1,3)-β-D-glucan (BDG) testing (**Supplementary Table 1**). Microbial cultures were performed using urine, blood, or sputum samples. Urine and sputum samples were inoculated onto appropriate culture media at 37°C for 24-48 hours. For blood cultures, samples were inoculated into both aerobic and anaerobic culture bottles and placed in an automated blood culture system (e.g., BACTEC, BacT/Alert) to detect bacterial growth, typically over a period of up to five days. Bacterial identification was carried out using the VITEK 2 Compact system (BioMérieux, France) according to the manufacturer’s instructions.

Suspected pathogen antigens and antibodies were detected using immunofluorescence assays (IFA), targeting seven pathogens: rotavirus, Influenza A, Influenza B, adenovirus, *Mycobacterium tuberculosis*, *Mycoplasma pneumoniae*, Epstein-Barr virus. The PCR tests targeted ten pathogens: *M. tuberculosis*, *M. pneumoniae*, Influenza A, Influenza B, Epstein-Barr virus, cytomegalovirus, respiratory syncytial virus, rhinovirus, and adenovirus. DNA extraction for PCR tests was carried out using extraction or purification reagents from Sansure Biotech Inc., following the manufacturer’s instructions. The *Cryptococcus neoformans* ink stain microscopy test was performed using fresh cerebrospinal fluid (CSF) samples, which were centrifuged at 3000 rpm for 15 minutes, and the sediment was used for smears. BDG testing was conducted using the commercial kit (Dynamiker Biotechnology (Tianjin) Co., Ltd.) to detect fungal cell walls, according to the kit’s instructions.

### 2.3 Nucleic acid extraction, library construction, and sequencing

Throat swabs were collected from the patients in tubes containing 1.5 ml of conservation liquid and thoroughly vortexed for 3 minutes. Subsequently, 1.3 ml of the homogenized sample was transferred to a new 1.5 ml centrifuge tube, and 13 µl of an external-internal reference was added. The sample was then centrifuged at 12,000 rpm for 5 minutes to enrich the pathogen nucleic acid. After centrifugation, 700 µl of the supernatant was discarded, and the remaining sample was mixed well. A 250 µl of the sample was processed using the MasterPure DNA&RNA Extraction Kit (KS118-BYTQ-24, KingCreate Biotech) for nucleic acid extraction and purification.

The synthesis of complementary DNA (cDNA) from RNA and the construction of targeted libraries were using the KM 50TM Plus Assay Kit (KingCreate Biotech) following the manufacturer’s instructions, which included fragmentation, end repair, target region enrichment, adapter ligation, and library amplification. The enrichment panel targeted 107 common respiratory tract pathogens, including viruses (31 DNA viruses and 38 RNA viruses), bacteria (10 Gram-positive bacteria and 12 Gram-negative bacteria), *mycoplasma* (1), and *chlamydia* (4) (**Supplementary Table 2**). The targeted pathogens had at least one completed genome sequence and had been previously reported in research studies. The probe design selected conserved regions such as ribosomal RNA genes (16S rRNA, 18S rRNA, or internal transcribed spacer), and housekeeping genes, allowing for pathogen identification down to the genus or species level. All libraries were sequenced on the KM MiniSeqDx-CN platform (KingCreate Biotech) using the paired-end model, with an average of 1 million reads conducted per sample. Sequencing was performed by KingMed Diagnostics (Hong Kong) Limited.

### 2.4 Bioinformatic analysis

To obtain high-quality reads, sequences with adapters, low quality, excessive N bases, and lengths below 35 bp were filtered using fastp (version 0.20.1) [25]. Only the samples with Q30 ≥75%, a clean read number ≥50K, and the external internal reference read number >200 were used for subsequent analysis. The clean reads were aligned to the human reference (hg38) using Bowtie2 (version 2.4.1) with the parameter “--very-sensitive” to remove the human reads [26]. Subsequently, the remaining reads were compared to the classification reference genome database, which contains all targeted pathogens and other species in their genus. This database also included species outside the genus with genome average nucleotide identity >80%. The species genomes were collected and downloaded from the National Center for Biotechnology Information (NCBI) genome database (ftp://ftp.ncbi.nlm.nih.gov/genomes/). The number of reads per million (RPM) at the species and genus levels was calculated. Bioinformatic analysis was performed by KingMed Diagnostics (Hong Kong) Limited.

### 2.5 Criteria for diagnosis decisions

The species in the databases were classified into four categories: Category A includes pathogenic species; Category B consists of opportunistic pathogens; Category C comprises normal respiratory microbiota; and Category D (others). Category A pathogens were considered positive if any sequence was detected. For Category B pathogens, the sequence number higher than 2000 was required to be considered positive. Similarly, for category C pathogens, the sequence number higher than 3500 was necessary to be deemed positive. The species in category D need physicians to confirm the authenticity based on clinical symptoms.

All samples were evaluated according to disease guidelines, clinical symptoms, CMT results, and tNGS panel results by at least two attending physicians. Given that the study included hospitalized patients with respiratory infections, any pathogen detected by either CMTs or tNGS was considered a true positive. Conversely, samples in which no pathogen was detected were regarded as false negatives. The treatment was adjusted according to the CMTs and tNGS diagnosis results.

### 2.6 Evaluation of Clinical Effectiveness of tNGS

The clinical effectiveness of tNGS was assessed based on the treatment response status of patients. At least two attending physicians reviewed the patient’s medical records to determine the impact of tNGS results on treatment plans. Adjustments to treatment plans were categorized as escalation, de-escalation, and no change (**Supplementary Table 3**). The impact of these adjustments on patient outcomes was further categorized as improvement, deterioration, or no change (**Supplementary Table 3**).

### 2.7 Statistical analysis

The sensitivity and specificity of CMTs and tNGS, determined based on whether the test reflected the clinical diagnosis, were compared using the Chi-square test online tool (https://www.shuxuele.com/data/chi-square-calculator.html). Data analysis was performed using SPSS software (version 21.0; IBM Corp., Armonk, NY, USA) and visualized using the ggplot2 package in R [27]. All tests were two-tailed, and statistical significance was set at P<0.05.

## 3. Results

### 3.1 Patient baseline characteristics

In this retrospective study, we collected data from pediatric patients diagnosed with respiratory tract infections (RTIs) at Qingdao Huangdao District People’s Hospital from April 1, 2023, to December 31, 2023. A total of 1209 pediatric patients with electronic medical records and targeted next-generation sequencing (tNGS) during this period. Of these, 269 patients were excluded due to multiple visits or lack of conventional pathogen detection records. Ultimately, 940 patients were included in the analysis.

Among the 940 patients included in the study, 785 (83.51%) were diagnosed with pneumonia, 86 (9.15%) with severe pneumonia, and the remaining patients with conditions such as tonsillitis and bronchitis (**Supplementary Table 4**). The average age of the patients was 4.83 years, with males accounting for 43.34% of the cohort. The average hospital stay was 6.14 days. Blood test results indicated that the average levels of procalcitonin and erythrocyte sedimentation rate were higher than the reference range, with 91.80% of the participants having elevated procalcitonin levels (**Table 1**).

**Table 1.** Demographic characteristics.

| Characteristics | Total number (%) | Average | Outlier (%) |
| --- | --- | --- | --- |
| Age (year) | 940(100) | 4.825 |  |
| Male | 445 (43.34) |  |  |
| Female | 495(52.66) |  |  |
| Length of hospital stay (day) | 940 | 6.14 |  |
| Blood laboratory examination (range) |  |  |  |
| Neutrophil number, 10<sup>9</sup>/L (2-7) | 442(47.02) | 3.88 | 165(37.33) |
| Monocyte number, 10<sup>9</sup>/L (0.12-0.8) | 442(47.02) | 0.56 | 65(14.71) |
| Basophil number, 10<sup>9</sup>/L (0-0.1) | 442(47.02) | 0.028 | 2(0.45) |
| Eosinophil number, 10<sup>9</sup>/L (0.05-0.5) | 442(47.02) | 0.11 | 204(46.15) |
| Lymphocyte, 10<sup>9</sup>/L (0.8-4) | 443(47.12) | 2.95 | 87(19.64) |
| Procalcitonin, ng/ml (0-0.05) | 841(89.47) | 0.30 | 772(91.80) |
| IgE IU/ml (0-100) | 590(62.77) | 201.13 | 263(44.58) |
| Erythrocyte sedimentation rate, mm/h (0-15/20) | 252(26.81) | 28.82 | 163(64.68) |

### 3.2 Diagnostic performance of CMTs and tNGS

All selected patients underwent both CMTs and tNGS diagnosis. The distribution of CMTs is shown in Supplementary Table 1. By comparing the diagnostic performance of CMTs and tNGS, we found that patients identified as infected by both methods accounted for 28.08%, with 22.35% of these samples detecting the exact same pathogens (**Figure 2, Supplementary Table 4**). However, 53.41% of the patients identified as infected showed completely different pathogens between the two methods. Patients identified as positive only by tNGS accounted for 63.3%, while those identified as positive only by CMTs accounted for 1.6%.

**Figure 2.**
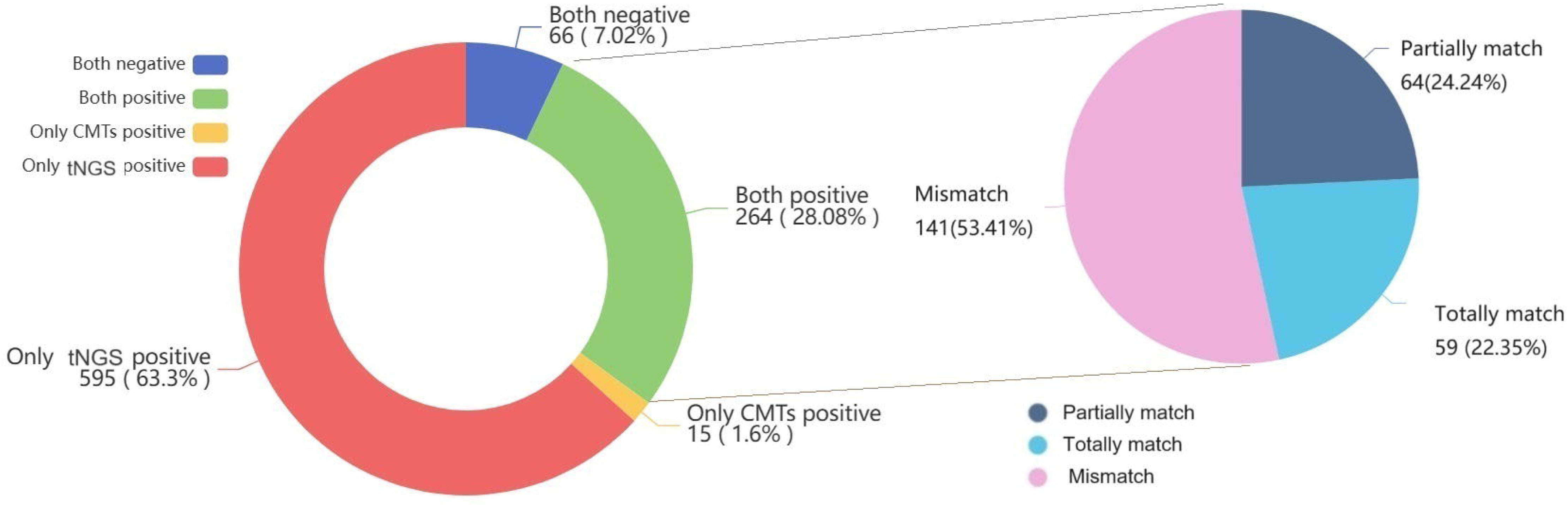
Consistency between CMTs and tNGS Results. In 940 patients with RTIs, 595 patients (63.3%) were CMTs negative and tNGS positive, and 15 patients (1.6%) were CMTs positive and tNGS negative. Sixty-six cases (7.02%) were negative for both CMTs and tNGS. Additionally, 264 patients (28.08%) were positive for both CMTs and tNGS.

At the pathogen level, tNGS detected a significantly higher variety of pathogens compared to CMTs in both viruses and bacteria (P < 0.05) (**Figure 3**). A total of 47 pathogens were identified and considered underlying pathogens. Among these, *Mycoplasma pneumoniae*, *Haemophilus influenzae*, and Epstein-Barr Virus (EBV) were detected by both methods in some patients, but tNGS still showed a significant advantage in detecting these pathogens (P < 0.05). *Enterococcus faecium* and *Mycobacterium tuberculosis* were exclusively detected by CMTs, which also identified untyped adenovirus strains in two patients and untyped rotavirus strains in four patients that were not detected by tNGS. For viruses, all RNA viruses were only detected by tNGS, which also has the capacity to identify virus sub-types.

**Figure 3.**
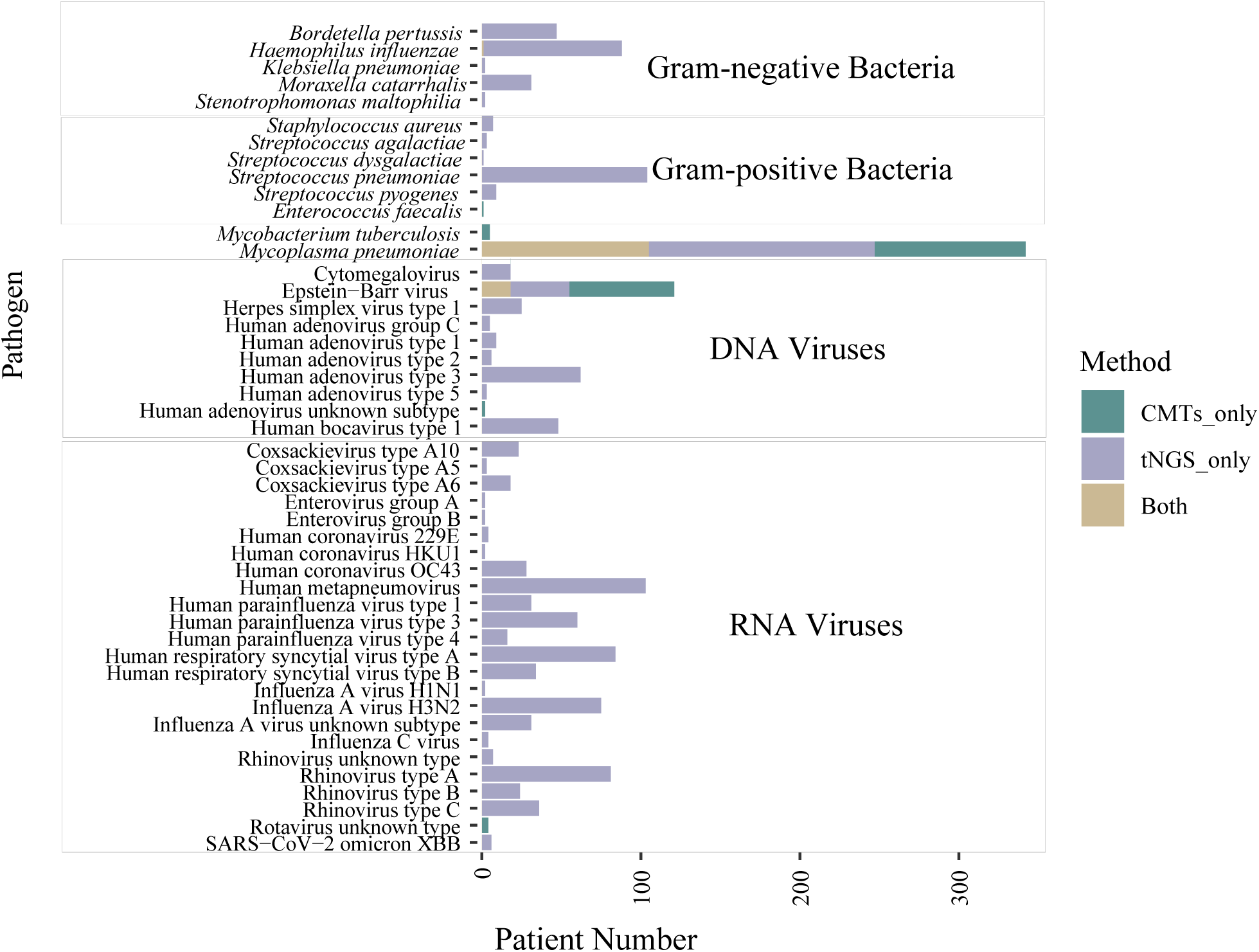
Detecting performance of CMTs and tNGS at the pathogen level. Specifically, a total of 47 pathogens were detected in all patients. tNGS showed significant advantages compared to CMTs in detecting both viruses and bacteria. The p-values were calculated using the Chi-square test, with *p<0.05 and ***p<0.001 indicating statistical significance.

The imaging data from the CT scans were also included in this study to compare the consistency of tNGS. The sensitivity of tNGS in diagnosing infections was 91.38%, and the sensitivity of imaging diagnosis was 82.45%, both of them significantly higher than the 29.68% sensitivity of CMTs (P < 0.001) (**Table 2**).

**Table 2.** Diagnostic sensitivity of the three methods.

|  | tNGS | CMTs | CT |
| --- | --- | --- | --- |
| True positive | 859 | 279 | 775 |
| False negative | 81 | 661 | 96 |
| Sensitivity | 91.38% | 29.68% | 88.98% |

### 3.3 tNGS-based treatment adjustment and clinical outcomes

Based on the tNGS diagnosis, the treatment of patients was adjusted through escalation, de-escalation, and no change. Escalation primarily involved the addition of medications and the extension of treatment duration, while de-escalation mainly involved the discontinuation of medications and dose reduction (**Supplementary Table 3**). A total of 332 patients (35.32%) had their treatment plans escalated, which included 193 cases with the addition of new medications, 49 cases with prolonged treatment duration, and 90 cases involving other interventions (**Figure 4**). Additionally, 273 patients (29.04%) had their treatments de-escalated, which included 166 cases with discontinued medications, 47 cases with reduced dosages, and 60 cases involving other adjustments. Furthermore, 335 patients (35.64%) experienced no change in their treatment plans.

**Figure 4.**
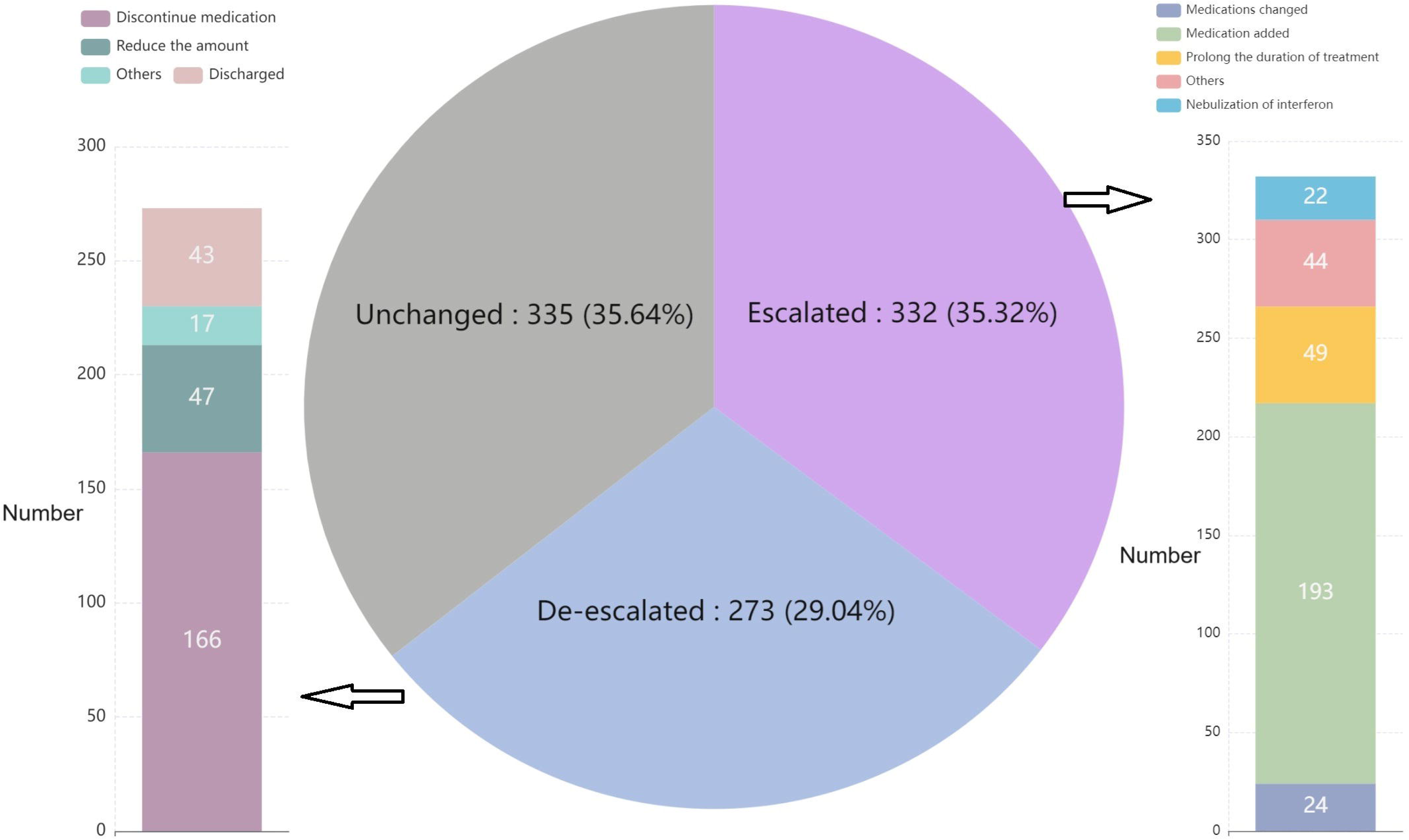
Distribution of detailed clinical adjustment strategies based on tNGS diagnosis.

After adjusting treatments guided by tNGS results, among the patients who had their treatment plans escalated, three experienced a deterioration in their condition,

while the rest showed improvement. All patients with de-escalated treatment plans showed improvement (**Supplementary Table 4,** **Figure 4**). The three patients who experienced deterioration were all infected with *M. pneumoniae*; one had refractory pneumonia; one had refractory pneumonia, and two had severe pneumonia (**Supplementary Table 4**). This data demonstrates that tNGS-guided treatment adjustments had a significant impact on patient outcomes, with most patients showing improvement regardless of whether their treatment was escalated or de-escalated.

### 3.4 Differences in pathogen infections across ages, genders, and seasons

To elucidate the influences on pathogen infections, a comparative analysis was conducted among children across different age groups, genders, and seasons (**Figure 5, Supplementary Table 5**). Twenty-eight pathogens showed variation among different age groups. Infections were predominantly concentrated in the 4-7 year age group and the 1-3 year age group, with 432 cases (49.59%) and 242 cases (27.78%), respectively. *Cytomegalovirus* (P<0.001) and Human respiratory syncytial virus type A (P<0.001) exhibited higher detection rates in children younger than one year. Human bocavirus type 1 (P<0.001) and Rhinovirus type A (P<0.001) were more prevalent in children aged 1-3 years. *Streptococcus pneumoniae* (P<0.001), Human metapneumovirus (P<0.001), and Rhinovirus type C (P<0.001) were more commonly detected in the 4-7 year age group. *M. pneumoniae* (P<0.001), Epstein-Barr virus (P<0.001), and Influenza A H3N2 (P<0.001) were significantly more prevalent in children older than seven years, with infection rates increasing with age. No significant differences were observed between gender groups, except for *H. influenzae*, which was slightly more prevalent in male children (P<0.05), and Human coronavirus 229E, which was more frequently detected in female children (P<0.05).

**Figure 5.**
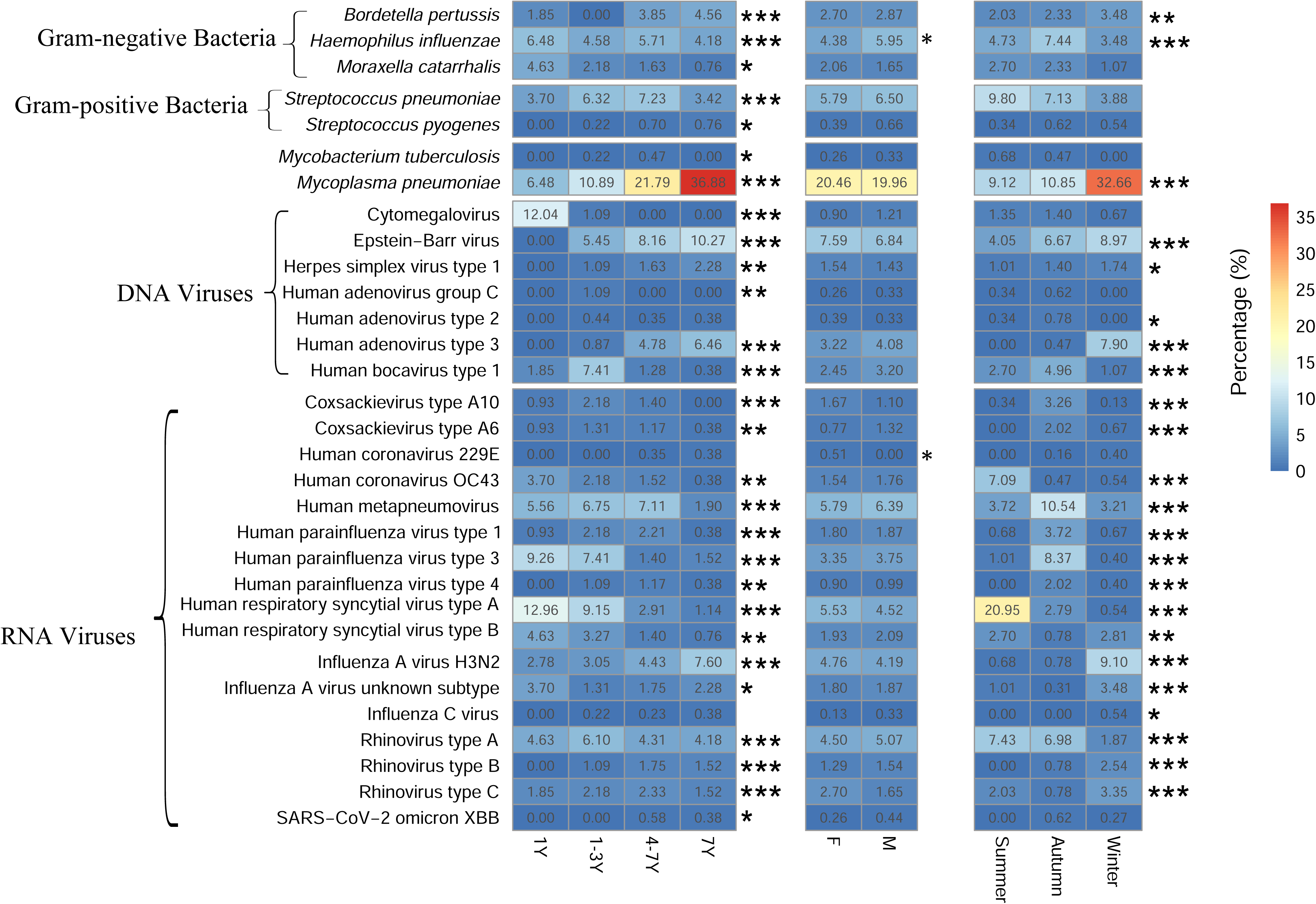
Comparison of pathogen detection in patients across different seasons, ages, and genders. The font color represents the group with the highest detection rate. * P < 0.05, ** P < 0.01, and *** P < 0.001.

The number of infected patients and the types of pathogens varied across different seasons. Our study detected 157 cases in summer, 320 cases in autumn, and 394 cases in winter, indicating a seasonal trend with infections gradually increasing from summer to winter. Different pathogens were observed to be prevalent during different seasons, with 18 pathogens showing significant variation (P<0.001). Human respiratory syncytial virus type A (P<0.001), Human coronavirus OC43 (P<0.001), and Rhinovirus type A (P<0.001) had significantly higher infection rates in summer. Conversely, *M. pneumoniae* (P<0.001), various Human adenovirus and Influenza viruses (P<0.001) had higher detection rates in winter. These seasonal trends demonstrate the influence of environmental factors on the transmission and prevalence of respiratory pathogens.

## 4. Discussions

Compared to adults, pediatric respiratory tract infections (RTIs) can have a prolonged impact, increasing the risk of wheezing, asthma, and reduced lung function in later life. Rapid pathogen discovery methods such as metagenomic next-generation sequencing (mNGS) and targeted next-generation sequencing (tNGS) have been developed to improve clinical diagnosis. However, few studies have focused on long-term and large-scale pediatric populations to evaluate the clinical performance and outcomes of these methods using specific sample types. In this study, tNGS to throat swab samples were used for diagnosing pediatric RTIs, we retrospectively evaluated its clinical utility and outcomes at a North China Hospital from April 1, 2023, to December 31, 2023. A total of 940 children with RTIs who were hospitalized in the pediatric department during this period were included in the study (**Figure 1**). Only patients with detailed CMTs recorders were included to provide a robust foundation for comparison with the tNGS diagnosis We observed that the blood tests of most patients showed abnormal values of procalcitonin (PCT, 91.8% in total) and erythrocyte sedimentation rate (ESR, 64.68% in total) (**Table 1**). PCT is unaffected by hormone levels in the body and has good stability, making it a useful marker for distinguishing between bacterial and non-bacterial infections and for evaluating the progression and prognosis of severe inflammatory diseases [28]. Studies have confirmed that elevated PCT levels are proportional to the severity of the infection, providing valuable guidance in predicting disease severity [29–31]. Although PCT is primarily used as an indicator of bacterial infections, our statistical data showed that PCT levels also increased during viral or mycoplasma infections, indicating that while PCT testing is highly sensitive, it may lack specificity. This could be due to the high expression of the *calc-1* gene in the thyroid during infection, which can result in increased serum PCT levels [32]. An accelerated ESR can be observed in various diseases such as pneumonia, active tuberculosis, rheumatic fever, severe anemia, and severe acute infections, making it a non-specific indicator of inflammation [33]. Although this study found that 64.68% of the children had elevated ESR, ESR alone is unable to be a sensitive and specific indicator for pediatric respiratory infections.

The mNGS has been applied in many RTI studies and has shown higher sensitivity than CMTs. Compared with mNGS, the probe capture-based tNGS method improves the proportion of target reads, processing speed, and cost-efficiency. In the present study, tNGS demonstrated significantly higher sensitivity (91.38%) in pathogen detection compared to CMTs (29.68%) (**Table 2**), exhibiting s similar performance to mNGS and surpassing CMTs [34, 35]. In terms of pathogens, tNGS was able to identify a broader range of pathogens, including low-abundance and RNA viruses, which are often missed by CMTs (**Figure 3**). Even for three pathogens detected by both two methods, tNGS also showed significantly higher sensitivity than CMTs (P < 0.05). Furthermore, tNGS often identified multiple infections, whereas CMTs typically detected a single pathogen, similar to findings from other studies [36, 37]. *Enterococcus faecium* and Rotavirus were only detected by CMTs because these two pathogens are not covered by our tNGS panel, indicating the limitations of tNGS in its only focus on targeted pathogens. For all targeted pathogens, our tNGS method showed similar diagnosis performance to mNGS and superior to CMTs, suggesting that more targeted pathogens should be included in this tNGS panel in the future.

This high sensitivity of tNGS is crucial for accurate diagnosis and supports the clinical utility in improving diagnostic accuracy and guiding appropriate treatment decisions in pediatric patients. After the implementation of tNGS testing, 35.32% of patients (332) had their treatment plans escalated (**Figure 4**). Among these, most patients showed improvement, and only three patients experienced deterioration, indicating that early and accurate identification of pathogens reduces the risk of severe disease progression. In patients who had their treatment plans de-escalated, none experienced deterioration, demonstrating that tNGS assists doctors in making precise adjustments and reducing the misuse of antibiotics, thereby preventing the development of drug resistance. For example, the incidence of refractory pneumonia with *M. pneumoniae* infection has increased due to drug resistance, resulting in adverse outcomes and sequelae [4, 38].

Respiratory pathogens often exhibit seasonal trends, which were also observed in our study. A previous epidemiological report on a child cohort in Guangzhou, a southern city in China, indicated that Human adenovirus was predominantly detected in summer [39]. In contrast, we observed higher proportions of Human adenovirus in autumn and winter, which may be due to the markedly different climates in these two regions. The overall seasonal trend for the Influenza virus in our study was consistent with findings from a long-term etiological and epidemiological study in China [40]. Given the ongoing development of children’s body functions and immune systems, they exhibit age-specific differences in pathogen infections. Numerous studies have emphasized the higher susceptibility of children, particularly infants and preschool-aged children, to acute respiratory infections [39–41]. Our study similarly found a high incidence of respiratory infections in preschool children aged 1-6 years. This distribution suggests age-specific susceptibility to different pathogens, potentially due to varying levels of immunity and exposure. Additionally, we didn’t find significant differences between different genders, which is consistent with previous studies [41].

## 5. Conclusions

In conclusion, our study confirms the advantages of tNGS over CMTs in diagnosing pediatric RTIs. The comprehensive detection capability of tNGS can significantly enhance clinical decision-making, leading to better patient outcomes.

Future research should focus on improving and standardizing tNGS methodologies to ensure accurate results across different clinical cases. The findings also emphasize the need for tailored diagnostic, treatment approaches, and preventions based on seasonal and age differences in pathogen prevalence.

## Author contributions

LXW, LXY, HZH, and XY designed the study and wrote and revised the manuscript. HZH and YF were involved in collecting, analyzing, or interpreting research data and writing the manuscript. LXW and LXY analyzed research data. All authors contributed to the article and approved the submitted version

## Institutional Review Board Statement

The study protocol was approved by the Ethics Committee of Qingdao Huangdao District People’s Hospital (LL-LW-2024001).

## Informed Consent Statement

Written informed consent was obtained from all adult participants or their legal guardians for participation in this study.

## Data Availability Statement

The targeted next-generation sequencing reads can be accessed in the China National GeneBank under BioProject accession number CNP0005922. All data will be publicly available after acceptance.

## Data Availability

https://db.cngb.org/cnsa/project/page/sub059464/view/

https://db.cngb.org/

## Acknowledgments

We would like to thank all patients and their families for sharing with us the results of the testing.

---

Supplementary Table 1. Statistic of CMTs.

Supplementary Table 2. Targeted pathogens in the tNGS panel.

Supplementary Table 3. Criteria of treatment plan adjustments and response based on tNGS diagnosis.

Supplementary Table 4. Clinical diagnosis, treatment, and outcomes for collected patients.

Supplementary Table 5. Statistic of P values across different comparison groups.

